# A2B-COVID: A method for evaluating potential SARS-CoV-2 transmission events

**DOI:** 10.1101/2020.10.26.20219642

**Authors:** Christopher J. R. Illingworth, William L. Hamilton, Chris Jackson, Ashley Popay, Luke Meredith, Charlotte J. Houldcroft, Myra Hosmillo, Aminu Jahun, Matthew Routledge, Ben Warne, Laura Caller, Sarah Caddy, Anna Yakovleva, Grant Hall, Fahad A. Khokhar, Theresa Feltwell, Malte L. Pinckert, Iliana Georgana, Yasmin Chaudhry, Martin Curran, Surendra Parmar, Dominic Sparkes, Lucy Rivett, Nick K. Jones, Sushmita Sridhar, Sally Forrest, Tom Dymond, Kayleigh Grainger, Chris Workman, Effrossyni Gkrania-Klotsas, Nicholas M. Brown, Michael P. Weekes, Stephen Baker, Sharon J. Peacock, Theodore Gouliouris, Ian Goodfellow, Daniela De Angelis, M. Estée Török

## Abstract

Identifying linked cases of infection is a key part of the public health response to viral infectious disease. Viral genome sequence data is of great value in this task, but requires careful analysis, and may need to be complemented by additional types of data. The Covid-19 pandemic has highlighted the urgent need for analytical methods which bring together sources of data to inform epidemiological investigations. We here describe A2B-COVID, an approach for the rapid identification of linked cases of coronavirus infection. Our method combines knowledge about infection dynamics, data describing the movements of individuals, and novel approaches to genome sequence data to assess whether or not cases of infection are consistent or inconsistent with linkage via transmission. We apply our method to analyse and compare data collected from two wards at Cambridge University Hospitals, showing qualitatively different patterns of linkage between cases on designated Covid-19 and non-Covid-19 wards. Our method is suitable for the rapid analysis of data from clinical or other potential outbreak settings.

## Introduction

Having emerged via zoonotic transfer in late 2019, the COVID-19 pandemic remains an ongoing public health priority [1,2]. Understanding the nature of viral transmission is a key factor in all strategies to prevent and control disease spread. The earliest stages of the outbreak were characterised by small, localised clusters of infection [3,4]. Identifying chains of inked cases remains crucial for containing disease spread [5,6], particularly in healthcare settings. Outbreaks in these settings, however, provide a particular challenge to the identification of linked cases as regular new introductions from the community have to be distinguished from potential cases of nosocomial transmission [7–9].

Viral genome sequencing provides one strategy for identifying clusters of transmission. The rapid evolution of viral populations leads to the accumulation over time of genetic differences which distinguish linked from unlinked cases [10]. A broad range of phylogenetic approaches for identifying linked infection clusters have previously been described [11–15]

With this background, a recent study of hospital-based COVID-19 infection used genome sequencing to identify potential clusters of infection. In this study, sets of individuals with identical viral genome sequences were often verified by a process of epidemiological follow-up to correspond to likely cases of nosocomial transmission [5]. Data from genome sequencing allowed feedback in real time to clinical, infection control and hospital management teams to inform their response.

Whilst genome sequence data is of great value for studying viral transmission, it has some drawbacks in the context of SARS-CoV-2. Given the recent emergence of SARS-CoV-2, and the low diversity of global viral sequences, finding identical sequences in different individuals does not necessarily imply a connection between those people. Furthermore, sequence data is not the only type of information that may be available. For example, studies of known SARS-CoV-2 transmission events have quantified the distribution of times between the onset of symptoms and the transmission of the virus, and of the subsequent time between infection and the onset of symptoms [16–19]. In addition the contribution of asymptomatic individuals to transmission is difficult to ascertain as they may not be sampled [20,21]. Such information has implications for the analysis of potential transmission events. Information about the location of individuals might also contribute to the identification or ruling out of connections; people who never physically interact cannot directly transmit the virus from one to another. Potential, therefore, exists for novel approaches in the identification of transmission clusters.

New methodologies have the potential to gain insights into viral sequence data. For example, noise in the process of viral sequencing can affect phylogenetic analyses which rely on finding identical sequences. Multiple studies have considered the problem of noise in genome sequence data, particularly with regard to identifying variant frequencies [22–25]. The potential for error in variant frequencies means that a viral consensus sequence is also stochastic, since it is generated from a potentially diverse viral population. Further, evolution would be expected to change viral sequences over time. Metrics which account for measurement error and viral evolution may be advantageous for identifying linked cases of infection.

A variety of studies have used short-read viral sequencing to evaluate the nature and likely direction of viral transmission. The ability of such data to capture the diversity of within-host viral populations has proved very valuable in the assessment of transmission events [26–29]. However, the low cost of data collection via nanopore-based methods has often outweighed the additional precision provided by Illumina technology [30,31]. Analyses which can utilise the consensus sequences provided by such methods have broader application, increasing their value.

Here, we describe a tool which implements a combined statistical and evolutionary framework to analyse genome sequence data with location data and knowledge of SARS-CoV-2 infection dynamics to rapidly identify clusters of infection. Our approach provides a rapid data analysis, outputting information in an easily interpretable format. We demonstrate its use with augmented sequence data from outbreaks on hospital wards, illustrating the value of different kinds of data in this context. While designed with clinical application in mind, the generality of the properties of viral transmission make our method applicable to any setting in which appropriate data has been collected from more than one individual. We hope that our approach will be of value in ongoing public health efforts to combat the COVID-19 pandemic.

## Results

Our method exploits data from genome sequencing alongside other information about individuals and COVID-19 infection. Given short periods of time between samples, the extent of measurement error in a viral sample may exceed the extent of evolutionary change in a population [32]. As a preliminary step therefore, we evaluated the extent of measurement error in our sequencing pipeline.

We examined cases among data collected from patients at the Cambridge University Hospitals National Health Service Foundation Trust (CUH) for which more than one viral sample was sequenced. A total of 136 such patients were identified, with between 2 and 9 (median 2) samples collected from each individual and 336 samples in total. Intervals between pairs of samples varied from 0 to 39 days. Each sample gave rise to a consensus sequence. We filtered the data to remove sequences with less than 90% coverage of the genome. Combining these data through regression, we inferred a mean error rate of approximately 0.207 nucleotide errors per sequence (S1 Figure). While small, this rate is significant. In our model, the expected time between symptoms being reported from individuals in a transmission pair is 5.7 days; the expected amount of sequence evolution within this time is not greater than the expected difference between two sequences resulting from noise (S2 Figure).

For each pair of individuals in a dataset we compared symptom onset, location, and genome sequence data with an underlying model of transmission, identifying whether or not the data were consistent with the null hypothesis that direct viral transmission occurred between the two individuals. Our model produces a simple output, stating that the data are either ‘consistent’ with the hypothesis of transmission (nominal p-value > 0.05), that transmission is ‘unlikely’ to have occurred (p-value < 0.01) or that the case is ‘borderline’ (p-value between 0.05 and 0.01).

We applied our model to data from two wards within CUH, which we term X and Y. Ward X was a ‘green’ ward, used for patients considered to be free from COVID-19 infection. By contrast, ward Y was a ‘red’ ward within the hospital, designated for the treatment of patients with COVID- 19 infection, on which multiple cases of infection in healthcare workers (HCWs) were identified. Information collected for these individuals included genome sequence data from viral swabs, dates of symptom onset and dates on which individuals were present on the wards in question.

Our method combines multiple types of data, using the information available to identify potentially linked cases of infection (Figure 1). We conducted two analyses of the data from wards X and Y, the first looking at the complete data collected for individuals, including dates of symptom onset, viral genomes and location data, and the second excluding location data, considering only times of symptom onset and the viral sequence data.

**Figure 1:**
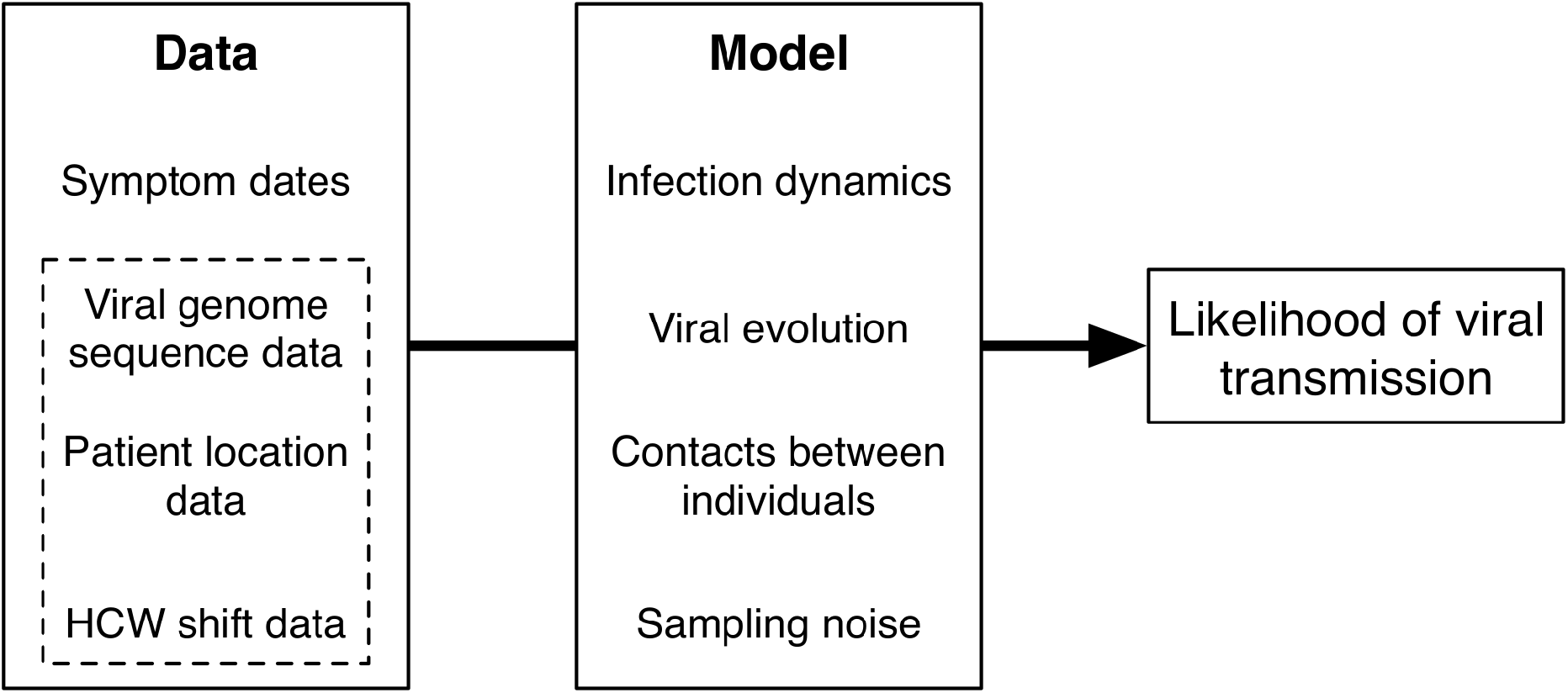
Overview of our method. Our approach estimates the likelihood that transmission could have occurred between pairs of individuals. The model takes as input dates on which individuals became symptomatic for COVID-19 infection. Further data which can be considered includes viral genome sequence data, and time-resolved location data for each individual. Our model combines details of COVID-19 infection dynamics with a model of viral evolution, information about potential contacts between individuals, and measurement error in the sequence data. Increasing amounts of data provide increasing amounts of resolution about the potential for viral transmission.

Analysing the complete data from each ward identified potential transmission events in each case (Figure 2). Outputs from our method are asymmetrical. For example, the data is consistent with transmission from 7069 to 7074 having occurred, but transmission from 7074 to 7069 was ruled unlikely. This is in part explained by 7069 reporting symptoms four days before 7074; the order of reporting symptoms provides information on the likely direction of transmission. Examination of the output from our code can be used to identify potential clusters of linked transmission events. For example, data from ward X suggest that the ten infections for which data were available were potentially mutually connected by transmission, though some underlying structure can be seen. The bottom three individuals coded 7108, 7128, and 7074, could each have infected each other, but have only borderline chances of having infected anyone else on the ward. These individuals could have been infected by either 7069 or 7112 infecting 7074, leading to further transmission events. The remainder of the individuals on the ward show multiple plausible transmission events, though we note that 7129 has only a borderline probability of having infected any other person. Examination of location data collected for the ward provides some insight into these findings (Figure 2B). Individuals 7108 and 7128 were never present on ward X, but were mutually in contact with 7074 throughout the duration of the outbreak via known household contacts. The health care worker 7129 was present on ward X at the same time as many of the other individuals, but became symptomatic at a later point; the later onset of symptoms suggests that 7129 was infected by another individual in the outbreak without passing the virus on within the ward. The overall picture we discover is what may be a single outbreak of linked cases on ward X, potentially started by a patient, but largely involving HCWs both on and off the ward.

**Figure 2:**
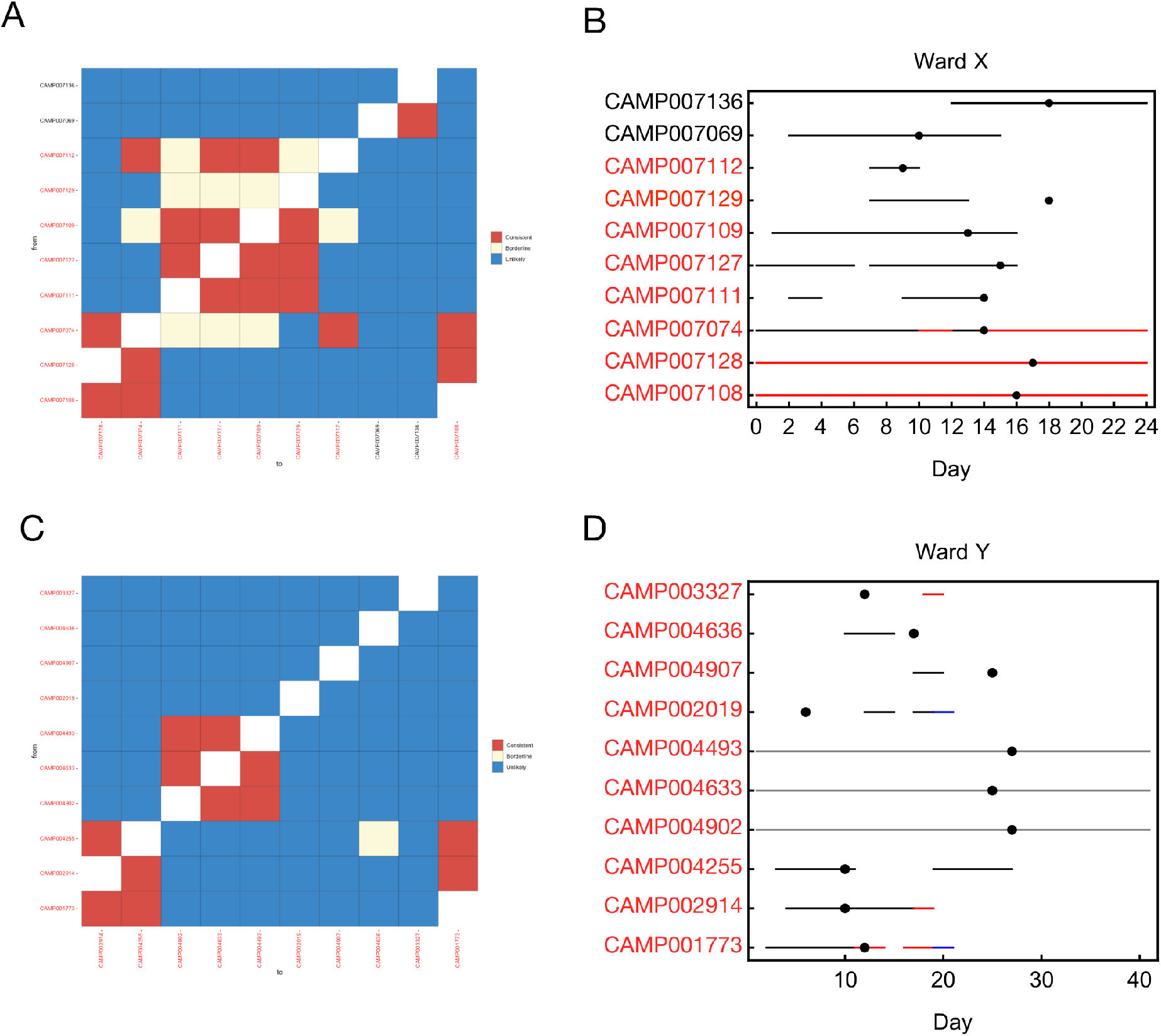
Analysis of the full datasets collected from wards X and Y. **A.** Output from the a2bcovid package given data from ward X. The plot shows potential links between cases, assessed in a pairwise fashion between potential donors (rows) and recipients (columns). Identifiers of individuals are coloured in either black (patients) or red (HCWs). Squares in the grid indicate that transmission from one individual to another is consistent with our model (red), borderline (yellow) or unlikely (blue). **B.** Locations of individuals linked to the ward X outbreak. Black lines indicate presence on ward X. Red lines indicate known household contacts between three individuals. Dots show times at which individuals first reported symptoms. **C.** Output from the a2bcovid package given data from ward Y. **D.** Locations of individuals linked to the ward Y outbreak. Black lines indicate presence on ward Y. Red and blue lines show presence in locations other than ward Y.

Analysis of data from ward Y revealed a less complex chain of events (Figure 2C). Precisely two clusters of individuals were inferred from the complete dataset, with individuals 1773, 2914, and 4255 forming one mutually interconnected set of infections, and the individuals 4902, 4633, and 4493 forming a second. While location data was missing for the latter three individuals the mutual connections between individuals can again be understood from the location data that do exist (Figure 2D). We note that in ward Y, cases were spread over a longer period of time than in ward X. In this case, we find two potential outbreaks between HCWs on the ward, with other cases reported on the ward not being linked to these individuals. Our results reflect the ‘green’ and ‘red’ natures of the two wards. Where ward X was designated for patients who were in theory free from COVID-19 infection, a coherent cluster of infection, potentially indicating a single introduction of the virus into the ward, was responsible for all of the observed cases. By contrast ward Y, being designated for COVID-19 cases, had multiple introductions of the virus onto the ward, perhaps two of those cases leading to ongoing nosocomial transmission.

Sensitivity analyses suggested that the measurement error has some effect on the outputs of our model. Calculations performed with increased and decreased error parameters led to changes in which events were identified as ‘Consistent’, ‘Borderline’ or ‘Unlikely’ (S3 Figure). Reducing the input error parameter to zero has the potential to induce more significant changes in our model output, as discussed further in the Methods section.

In order to assess the value of location data we repeated our estimation in their absence, using only viral genome sequence data and the dates on which individuals first reported symptoms. Location data generally reduces the inferred potential for viral transmission; under the logic of our method, individuals who are not in the same place at the same time cannot infect one another. The value of location data in increasing the precision with which networks of links between cases of infection is shown by the results from ward X (Figure 3A). While the overall pattern of data shows multiple potential connections between individuals, the independence of the transmissions between 7108, 7128, and 7074 from the remainder of the network was lost in the absence of spatial data. Spatial data in our model was defined in terms of presence or absence on a ward, more refined information not being available. An analysis of data from ward Y showed an intriguing result, with previously unseen potential links between individual 2019 and the first cluster, and between 3327 and the second cluster (Figure 3B). These HCW displayed symptoms earlier than the people in their respective clusters, consistent with being the original cases in each case. Further, phylogenetic reconstruction showed that the sequences from these individuals were consistent with their being linked (Figure 3C; similar data for ward X is shown in S4 Figure). However, the location data do not show them as working shifts on ward Y at the same time as any of the other linked individuals were present. We suggest that unrecorded contacts may exist in this case.

**Figure 3:**
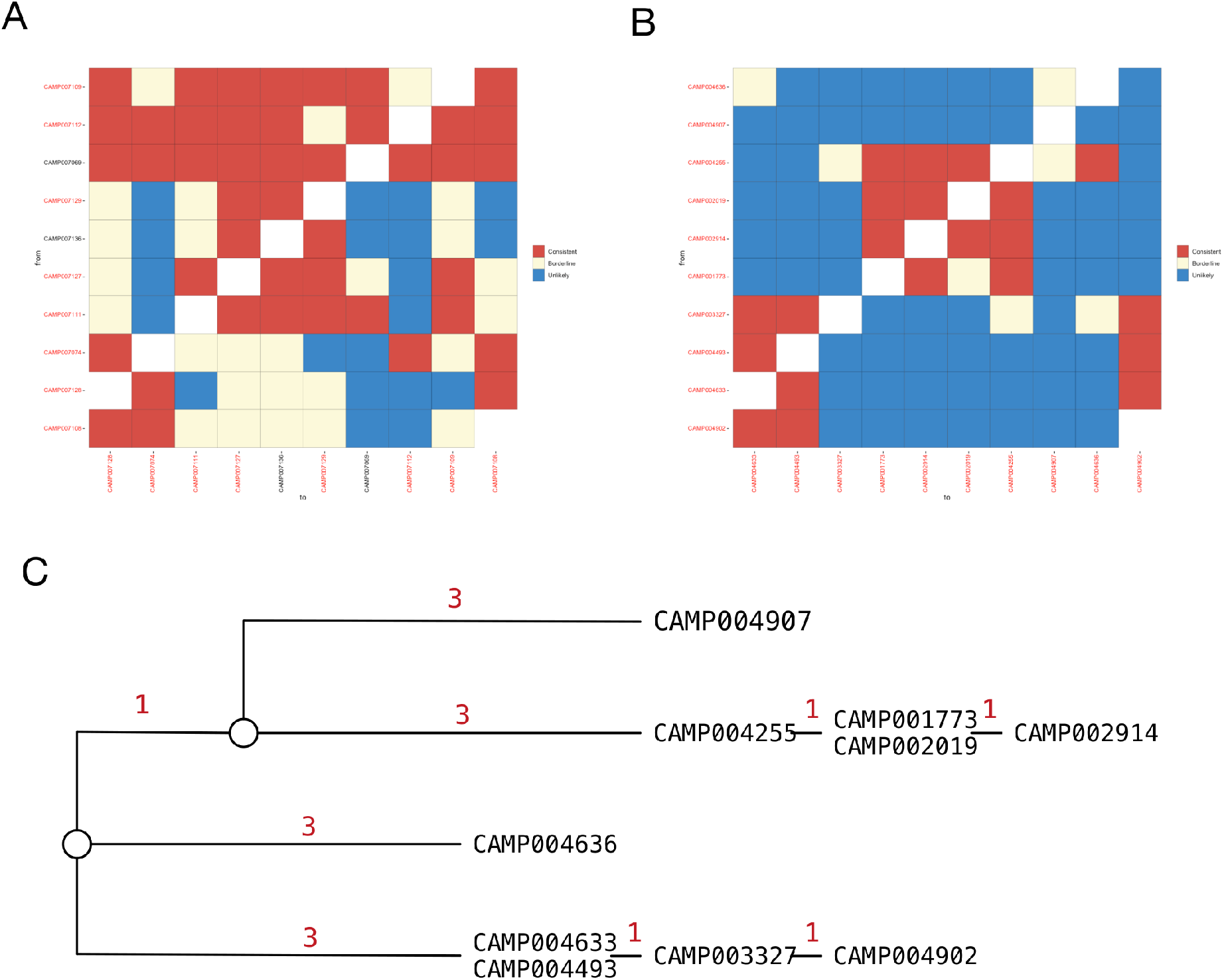
Analysis of the symptom onset and sequence data collected from wards X and Y. **A.** Output from the a2bcovid package given data from ward X, omitting location data for individuals. The plot shows potential links between infections. Identifiers of individuals are coloured in either black (patients) or red (HCWs). Squares in the grid indicate that transmission from one individual to another is consistent with our model (red), borderline (yellow) or unlikely (blue). **B.** Output from the a2bcovid package given data from ward Y, omitting location data for individuals. **C.** Phylogenetic relationship between sequences collected from individuals on ward Y. The tree was constructed manually using a principle of maximum parsimony[38]. Red digits indicate the number of substitutions between sequences from each individual.

The potential for missing location data to prevent the identification of linked individuals shows that measuring location can be difficult, more so for HCWs than patients. While patients are unlikely to be highly mobile, HCWs move around the hospital outside of their shifts. Unless explicitly recorded, off-ward contacts between HCWs are unlikely to be noted, leading to the potential non-inference of genuine links. While missing location data cannot be unambiguously diagnosed as the cause of our result, this case highlights the limitations intrinsic to our approach. Our software may provide valuable insights, but does not replace the need for full epidemiological investigation.

## Discussion

We have here set out a method for identifying potential cases of direct transmission between pairs of individuals, based upon the dynamics of SARS-CoV-2 infection, data describing times of co-location between individuals, and genome sequence data collected during infection. In a first application of our method, we analysed data from two hospital wards. In each, we identified cases where the data were consistent with viral transmission occurring between either patients or HCWs on the ward. Our method builds upon information that can be obtained from a phylogenetic analysis, incorporating data from multiple sources to present an easily-interpretable map of potential linked cases of infection. It is likely to be valuable in the initial assessment of potential cases of nosocomial transmission, highlighting pairs or clusters of individuals for further epidemiological assessment, and allowing for a more strategic deployment of resources for outbreak investigation and targeted interventions.

Our method brings together a variety of data, combining an evolutionary model for the analysis of sequence data with location information and details of the dynamics of viral infection. In contrast to standard phylogenetic approaches to sequence data, our model explicitly accounts for noise in the generation of a viral consensus sequence; using within-host data we identified a magnitude of error of a fraction of one nucleotide per genome. In rapidly evolving viruses for which transmissions are separated by longer periods of time, the within-host evolution of viral populations is likely to overwhelm the effect of noise in the sequencing process. However, for cases of acute infection, separated by only a few days, the extent of noise may be close to the expected evolutionary change in the population, making it an important factor to consider.

One limitation of our method is that it deals with consensus viral sequences rather than deep sequence data. Where available, detailed measurements of within-host viral diversity may lead to an improved picture of relationships between cases of viral infection. We note further that our tool analyses data in a pairwise manner; while distinguishing plausible from implausible links between cases of infection, it does not attempt to infer a complete reconstruction of a transmission network. Unobserved cases of infection are not considered. Our model used parameters which in some cases have been derived from early studies into SARS-CoV-2 spread. To account for the event that further research leads to a better understanding of viral transmission we provide options to perform calculations with user-specified parameters. We finally note that a statistical inference from our model does not describe the probability of transmission having occurred between two individuals. Instead it describes how consistent the data are with transmission. Our model is intended as a first step towards further epidemiological investigation.

Our model has a range of features specifically tailoring it to the real-time analysis of data in a hospital context during an outbreak of a rapidly spreading viral disease. Our method is designed for simplicity both in being easy to use and in rapidly producing an interpretable output. To this extent our method is limited in what we try to infer, highlighting only pairs of individuals where the data are consistent with transmission. For example, in a case where an individual A infects B and C, it could be that our method highlights not only the real transmission events, but also reports that the data from B and C is consistent with transmission occurring between them. This does not comprise an error in our method, but does require our method to be understood. We note that, in a hospital environment, a positive output from our method could be followed up by investigative efforts and epidemiological follow up; such efforts have the potential to collect data beyond that considered by our method.

We believe that the key application of our method will be in investigating nosocomial transmission of SARS-CoV-2. Within a hospital, potential cases of transmission may be obscured by a large number of cases of community-acquired infection. In a busy clinical setting, our tool has the ability to rapidly separate potentially linked cases from those which are likely to be unlinked. In this way we allow investigative efforts and epidemiological followup to be focused more precisely, concentrating effort on cases where transmission is a real possibility.

## Methods

### Model overview

We here consider pairs of individuals, who for the purpose of notation, we describe as individuals A and B. Given data on when the individuals became symptomatic for SARS-CoV-2 infection, their locations, and their viral genome sequences, we generate a statistic to test whether the data are consistent with viral transmission having occurred from A to B.

To outline this process, suppose that we have observed data y from this pair of individuals. The null hypothesis of transmission is supported by the data if these data have high probability of having arisen given transmission from A to B. More formally, the hypothesis is accepted at a confidence level ψ if the probability of observing y, or data that are “less extreme” (i.e. data that are even more consistent with transmission than y) is at least ψ under the hypothesis of direct transmission from A to B. We outline our method in detail below.

### Available data

#### Notation

An overview of the notation used in the description of our model is shown in Figure 4. The dates of symptom onset and the dates when viral sequence data were collected are denoted S_A_ and S_B_ and D_A_ and D_B_, respectively. These dates are assumed to be known, or in the case of symptom dates can be estimated from times at which individuals tested positive. Further data described the locations of the individuals A and B on each day, with the binary indicator C_A_(L,T) denoting whether individual A was present in location L on day T. The information describing the location of individuals may be uncertain, so we represent it by w_A_(L,T), the probability that individual A is present in location L on day T. For example, if A is known to be in location L on day T we have w_A_(L,T)=1, while if A is known not to be in location L on day T we have w_A_(L,T)=0. If the location of A at this time is unknown, w_A_(L,T) is defined as described below. Analogously to this, the binary indicator C_AB_(T) denotes whether or not A and B were in contact on day T. Uncertainty in this indicator is represented by the probability w_AB_(T) that A and B were present in the same location on this day. In describing genomic data, H_A_ and H_B_ describe Hamming distances between the viral sequences collected from A and B and their mutual consensus. The CT scores of the viral samples are denoted V_A_ and V_B_.

**Figure 4:**
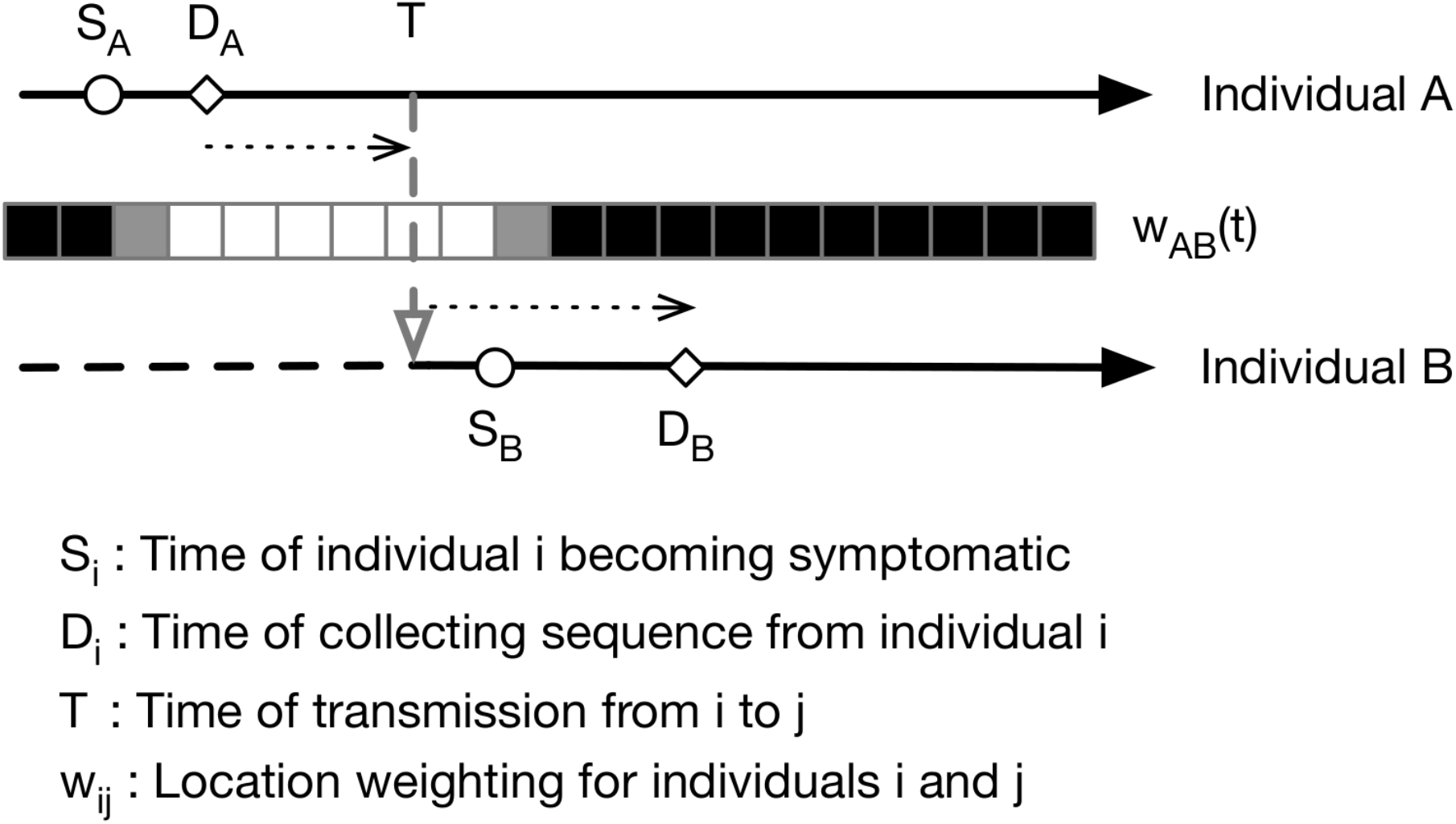
Notation used in our method. An overview of our model for transmission events is shown in Figure 3. We divide time into discrete days. For the individual A, we denote by S_A_ the date at which that individual became symptomatic, and by D_A_ the date at which a sample of viruses were collected for genome sequencing. For each pair of individuals A and B we denote by w_AB_(t) the probability that A and B were co-located on day t. Within our model, we assume that dates of sample collection are known, while times of symptom onset are known or estimated. Using these statistics, in combination with viral sequence data, we calculate a statistic describing the potential for individual A to infect individual B on any given day T. Summing this statistic across T, we obtain an estimate of the consistency of our data with transmission having occurred between the two individuals.

#### Symptom data

Due to extensive monitoring of individuals in hospital, we often had information on the dates of symptom onset for individuals. When these were unknown we estimated them from positive test dates. To perform this estimation we used symptom onset dates and positive test dates from 86 health care workers and 393 patients from Cambridge University Hospitals, fitting an offset gamma distribution to these data (S5 Figure, S1 Table). Where symptom dates were missing, the mean of this distribution was used to impute symptom onset dates from positive test dates. We write Ŝ_A_ to denote an estimate for S_A_. Where positive test dates are used in place of symptom onset dates, greater care is required in the interpretation of results.

#### Location data

In our study, time was measured in whole days. For example, if an individual was known to be in location L for any part of day T, we set w_A_(L,T)=1. Known location data were edited for health care workers to account for their increased mobility, night shifts which span more than one day, and uncertainties such as the potential for fomite transmission. If for a healthcare worker we had that w_A_(L,T)=1 for some L and T we set w_A_(L,T-1) and w_A_(L,T+1) to be equal to a minimum value of 0.5.

Where location data were missing it was necessary to specify values w_A_(L,T). Data from our study were centred on cases from a specific part of the hospital, usually a single ward; this location was denoted L*. Where location data were missing for a patient, we set w_A_(L*,T)=1 for all T, assuming that a patient was always on the most common ward. Where location data were missing for health care workers, we set w_A_(L*,T)=4/7 for all T, reflecting shift patterns. We note that in other circumstances (e.g. a dataset spanning an entire hospital), an alternative prior for the location of individuals could be more appropriate.

Contact information was derived from the location data. For any two individuals we note that there could be multiple locations in which they could be in contact on a single day. We combined probabilities of contact across potential locations, calculating

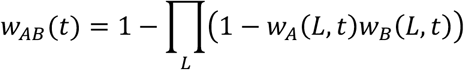

#### Viral genome sequence data

Consensus genome sequences were calculated from viral sequence data. Sequences were subjected to two levels of quality control. The first considered the coverage of the genome. An unambiguous nucleotide is here defined as an instance in which sequencing describes an A, C, G, or T. We applied the criterion that sequences had to unambiguously describe nucleotides at 80% or more of the sites in the genome.

The second level of quality control counted ambiguous nucleotides that were found at sites in the genome that were found to be polymorphic between the collected viral sequences. These sites are more likely to be informative with regards to the number of genetic differences between two sequences; a genome with high overall coverage but ambiguity at multiple of these positions would in practice be quite uninformative. Having identified polymorphic sites, we required sequences to have no more than one ambiguous nucleotide at these positions.

In some cases, multiple viral samples were collected from the same individual. Viral genomes collected from the same individual were usually extremely similar to one another (S1 Figure). In such a case, we identified the earliest sequence with sufficient coverage of the viral genome, using this sequence for analysis. Where positions in this genome were ambiguous, and where other sequences from the same individual had unambiguous nucleotides at these positions, the other sequences were used to construct a more complete consensus sequence for the individual.

Given viral sequences from the pair of individuals A and B we calculated Hamming distances from each sequence to a pairwise consensus sequence; we denote these distances as H_A_ and H_B_.

### Assessing viral transmission

We denote as X_T_ an indicator for the event that transmission took place at time T, and as X an indicator for the event that transmission took place at all. To test the hypothesis of transmission, we calculated the probability of observing the data y under the null hypothesis that transmission occurred, p(y|X) = Σ_T_ p(y|X_T_)P(X_T_|X), where P(X_T_|X) is the probability that transmission took place at T given transmission, which we abbreviate as P(T).

Let Y represent the observable data. Y consists of the symptom time S_B_, the Hamming distances H_A_ and H_B_, and the set of C_AB_(T) for all T, denoted C_AB_. We will write an expression for the probability of the observable data as follows:

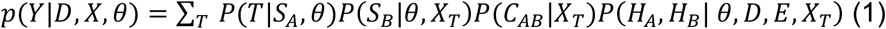

where D={D_A_, D_B_}, E is the error in sequencing, and θ represents the set of parameters that are assumed to be known. We note that we condition on S_A_; an alternative approach would be to write the equation in terms of S_B_-S_A_. We consider the parts of this equation in turn.

### Assessing viral transmission: Symptom and location data

In equation (1), P(T|S_A_,θ) describes the probability that transmission is at time T, where time is measured relative to S_A_, the time of onset of symptoms in A. This term describes the infectivity profile of the virus, that is, the time from symptom onset to transmission. We follow previously published work which has characterised this as an offset gamma distribution[16,17,35].

The term P(S_B_|θ,X_T_) describes the probability that B becomes symptomatic at time S_B_, given that transmission occurs at time T. Again we have information from the same literature characterising this as a lognormal distribution. We therefore write:

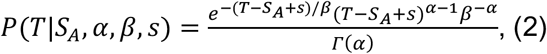

where s is the offset and α=97.1875, β=0.2689, and s=25.625, and

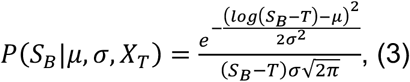

where μ=1.434, and σ=0.6612. Each of these expressions treat T as a continuous variable; we use an approximation to discretise the formula to a resolution of single days, obtaining

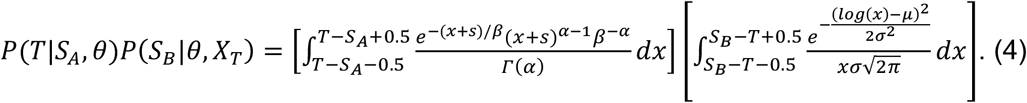

We next consider the term P(C_AB_|X_T_). Where |C| is the length of the vector C_AB_, we note that there are 2^|C|^ possible such vectors. If C_AB_(T)=0, transmission cannot have occurred at time T, so that, if transmission occurred at time, then C_AB_(T), the Tth element of C_AB_, equals 1. Regarding other elements of C_AB_, we take a naive approach to contact patterns, assuming that P(C_AB_(t)=1|X_T_)=0.5 for each t not equal to T. This implies the result that P(C_AB_|X_T_)=0.5^|C|-1^ for any C_AB_ with C_AB_(T)=1, with P(C_AB_|X_T_)=0 otherwise.

### Assessing viral transmission: Viral sequence data

Finally, we consider the term P(H_A_,H_B_|θ,D,X_T_), which is derived from the viral genome sequence data. In order to generate H_A_ and H_B_, we first calculated a local consensus sequence across all of the viral genomes in our data. Next, for each pair of sequences from individuals A and B, we calculated a pairwise consensus, defined as the nucleotide shared by the two sequences where the sequences agreed, and the nucleotide in the local consensus where the sequences differed. H_A_ and H_B_ were then calculated as the Hamming distances from each of the two sequences to the pairwise consensus sequence. These distances describe the number of substitutions observed to have been gained by the viral population in each individual.

In our analysis we assumed an infinite sites model; among our sequences any given mutation can be obtained only once, while the reversion of mutations back to the consensus never occurs.

We used a Poisson model to compare the number of observed substitutions in each sequence with an expected rate of viral evolution. Our model includes a term accounting for errors in the viral consensus sequences. In the notation of Figure 4, we note that if D_A_ is before T, any variants observed in sequence data from A but not in the data from B can only arise from error. Under our infinite sites assumption, such variants cannot revert in the time between D_A_ and D_B_ so must be caused by error in the observation. However, if D_A_ is after T, such variants have the potential to evolve in the time between D_A_ and T, in addition to being potentially caused by measurement error.

Similarly, variants observed in data from B but not from A can arise either from error, or as a result of evolution going back an assumed common ancestor at the earlier of the time of transmission T and the previous time of sequencing D_A_. We can thus describe the probability of observing the data H_A_ and H_B_ under the assumption of transmission at time T:

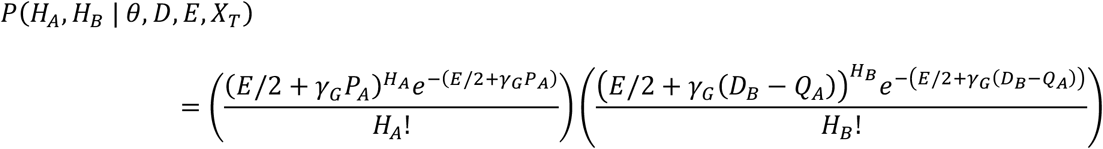

where P_A_ = max{0, D_A_ - T} and Q_A_ = min{D_A_, T}. The rate of evolution γ_G_ describes the expected number of substitutions per genome per day, while the parameter E is the mean number of errors in the Hamming distance between two viral sequences, estimated as described below.

### Estimating noise in genome sequence data

In order to estimate the extent of measurement error in a consensus viral genome, we examined cases among data collected at Cambridge University Hospitals (CUH) for which more than one viral sample was sequenced. We identified 136 such patients, with between 2 and 9 samples collected from each individual and 336 samples in total. Each sample gave rise to a consensus sequence; we filtered the data to remove sequences with less than 90% coverage of the genome. For each pair of samples i and j, collected from the same individual, we recorded H_ij_, the Hamming distance between them, ΔT_ij_, the absolute difference in time between the dates on which the samples were collected, measured in days, and the viral load of each sample, as represented by the CT scores V_i_ and V_j_.

Following in principle a previous approach to estimating noise and rates of evolution [32], we then fitted a Poisson model to the data, deriving for each pair the log likelihood

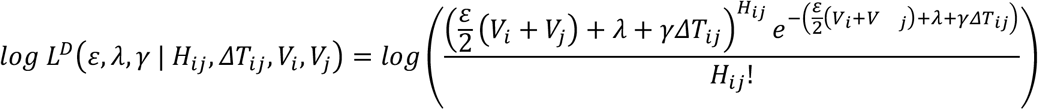

and estimating the parameters ε, λ and γ so as to maximise the sum of the log likelihoods across all pairs of sequences; we inferred the parameters 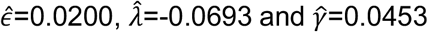. Here the value 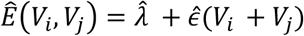 provides a simple estimate of the extent of measurement error in a Hamming distance, expressed in terms of the CT scores of the two samples. For the purposes of our model this function was evaluated at the mean CT score of 24.091. This provided an estimate for the pairwise difference arising through measurement error, 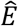, of 0.414 nucleotides, equivalent to 0.207 nucleotide errors per genome sequence. The estimate 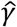 describes the mean rate of within-host evolution calculated across the within-host sample. It is expressed as a number of substitutions per genome per day, and is equivalent to a rate of 6.0 × 10^−4^ substitutions per locus per year, close to the value of 8 × 10^−4^ that has been calculated from global sequence data [33]. In so far as we require an estimated rate of evolution spanning both within-host and between-host evolution, we used in our model a rate 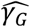 of 0.0655 nucleotides per day, equivalent to this latter, globally estimated, rate of evolution.

To examine the effect of CT score upon our inference, a repeat calculation was performed in which these data were ignored; this gave a worse fit to the data under the Bayesian Information Criterion[34] (S2 Table).

In a case where no sequence data was observed for an individual, we excluded that individual from our calculation. An option within our method allows for calculations to be performed between individuals where no sequence data was collected; under this option we set P(H_A_, H_B_|θ, D, E, X_T_)=1 for all A and B.

### Assessing viral transmission: Hypothesis testing

Having derived the expression (1) for P(Y|D,X), we now derive the probability P(y|D,X) of the specific observed data y. The data y consist of the symptom time S_B_, if it is known, the Hamming distances H_A_ and H_B_, the set of those C_AB_(T) that are known, and the information about potential locations and contacts in cases where the C_AB_(T) are unknown, which are encapsulated in w_AB_(T). p(y|D,X) is defined by setting Y to equal the data y that are observed, and then integrating P(Y|D,X) over the potential values for any missing data.

Integration was required with respect to the unknown contact dates, applying to the term P(C_AB_|X_T_). We generalise the argument made for this term in the case of Y to show that in this case we have

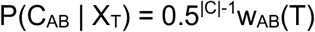

A full derivation is given in S1 Text.

We thus have the result

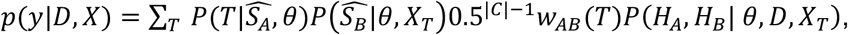

where θ={α,β,s,μ,σ,Ê,γ_G_}, and seek to compare this to potential values p(Y|D,X). To achieve this, for confidence level ψ, we define a threshold p_ψ_(D) by

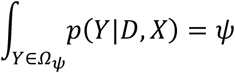

where Ω_ψ_={Y: p(Y|D,X) ≥ p_ψ_(D)}. Note that the threshold p_ψ_(D) depends on the sample collection times D={D_A_,D_B_}. For values ψ=0.95 and ψ=0.99, the observed data y is deemed ‘consistent’ with transmission if p(y|D,X) ≥ p_95_(D), ‘borderline’ if p_95_(D) > p(y|D,X) ≥ p_99_(D), and unlikely if p_99_(D) > p(y|D,X).

To identify these threshold values we calculated p(Y|D,X) across large numbers of sets of data Y, in which we assumed without loss of generality that S_A_=0. Calculations were performed for all Y in which S_B_ ∈ [-11, 87], and for all values H_A_ and H_B_ for which H_A_+H_B_ ∈ [0, 10] and H_A_ ∈ [0, H_A_+H_B_]; these ranges were chosen to return values of at least 10^−6^ from each component of p(Y|D,X). In our code these statistics are calculated for D_A_ ∈ [-10, 40], and D_B_ ∈ [S_B_-10, S_B_+40]; values outside of these parameters are unlikely.

In the integral, we note that there are a large number of possible vectors C_AB_ that indicate all times when a pair were in contact. We approximated the sum by generating 100 random vectors C_AB_ for each set of other parameters, and calculating the sum over these vectors, altering the value 0.5^|C|-1^ in P(C_AB_|X_T_) so as to normalise the integral. Reflecting our approach to contact patterns, we generated the C_AB_ as random vectors of draws from a Bernoulli distribution with mean 0.5. Repeating this calculation with different sets of 100 vectors did not substantially change the thresholds obtained. Our code allows for the generation of alternative thresholds with different probabilities of an element of C_AB_ being equal to 1. We note that if this probability is higher, fewer datasets will be judged consistent with transmission.

#### Study setting, participants and data collection

This study was conducted at Cambridge University Hospitals NHS Foundation Trust (CUH), a secondary and tertiary referral centre in the East of England. SARS-CoV-2 positive cases tested at the on-site Public Health England (PHE) Clinical Microbiology and Public Health Laboratory (CMPHL) were identified prospectively from 26th February to 17th June 2020. The CMPHL tests SARS-CoV-2 samples submitted from over thirty organisations across the East of England (EoE) region and all samples from CUH. The majority of samples were tested using an in-house validated qRT-PCR assay targeting the SARS-CoV-2 RdRp genes, as described in a previous publication [5], with more recent samples tested using the Hologic Panther™ platform [36]. Patient metadata were accessed via the electronic healthcare record system (Epic Systems, Verona, WI, USA). Metadata collected included patient demographic information, duration of symptoms, sample collection date and location (ward and hospital). Patients and samples were assigned unique anonymised study codes. Metadata manipulations were performed using the R programming language and the *tidyverse* packages installed on CUH Trust computers.

#### Sample sequencing

All samples collected at CUH and a randomised selection of samples from the EoE region were selected for nanopore sequencing on-site in the Division of Virology, Department of Pathology, University of Cambridge. This enabled us to rapidly investigate suspected hospital acquired infections at CUH as previously described [5]. Briefly, a multiplex PCR based approach was used according to the modified ARTIC version 2 protocol with version 3 primer set, and amplicon libraries sequenced using MinION flow cells version 9.4.1 (Oxford Nanopore Technologies, Oxford, UK). Sequences were made publicly available as part of COG-UK (https://www.cogconsortium.uk/) via weekly uploads with linked metadata onto the MRC-CLIMB server (https://www.climb.ac.uk/).

Samples collected via the CUH healthcare worker (HCW) screening programme were also prioritised for on-site nanopore sequencing, as previously described [37]. This programme entailed asymptomatic screening of selected wards, symptomatic testing of self-presenting HCW and testing of symptomatic contacts of positive HCW. After a HCW tested positive, members of the HCW screening team contacted the HCW and retrospectively collected data on symptom onset date, symptomatology, household contacts, their job role, and which wards they had worked in for the preceding two weeks. Most positive HCW could identify symptoms on retrospective questioning, even if they were identified in the asymptomatic screening arm; however, a small minority were genuinely asymptomatic and never went on to develop symptoms. HCW presenting acutely to medical services at CUH were not part of the HCW screening programme, but were identified as HCW from their medical records as part of hospital surveillance.

#### Identifying hospital-associated outbreaks for investigation

Patients tested at CUH were categorised on the basis of time between admission and first positive swab into different groups reflecting the likelihood that their infection was community or hospital acquired, as previously described (Meredith et al, LID 2020). The categories used were: 1) Community onset, community associated (first positive sample <48 hours from admission and no healthcare contact in the preceding 14 days); 2) Community onset, suspected healthcare associated (first positive sample <48 hours from admission with healthcare contact in the preceding 14 days); 3) Hospital onset, indeterminate healthcare associated (first positive sample 48 hours to 7 days post admission); 4) Hospital onset, suspected healthcare associated (first positive sample 8 to 14 days post admission); 5) Hospital onset, healthcare associated (first positive sample >14 days post admission); 6) HCW.

All CUH patients in categories 3, 4, and 5 (hospital onset with indeterminate, suspected or definite healthcare associated COVID-19 infections) and 6 (HCW) were included for analysis and integrated into the HCW screening dataset of positive HCW. The main wards the HCW had worked in prior to testing positive and the ward where each patient had first tested positive were used to identify ward clusters of hospital-associated infections. The ward clusters are named anonymously here as Wards X and Y.

### Ethics statement

This study was conducted as part of surveillance for COVID-19 infections under the auspices of Section 251 of the NHS Act 2006. It therefore did not require individual patient consent or ethical approval. The COG-UK study protocol was approved by the Public Health England Research Ethics Governance Group (reference: R&D NR0195).

## Data Availability

Sequence data analysed were generated by previous research studies and are available from the GISAID server. A list of sequence IDs are provided in Supplementary Material. Sequences and accompanying metadata has been uploaded onto the MRC-CLIMB server.

https://www.gisaid.org

https://www.climb.ac.uk/

## Supporting Information

**S1 Figure:**
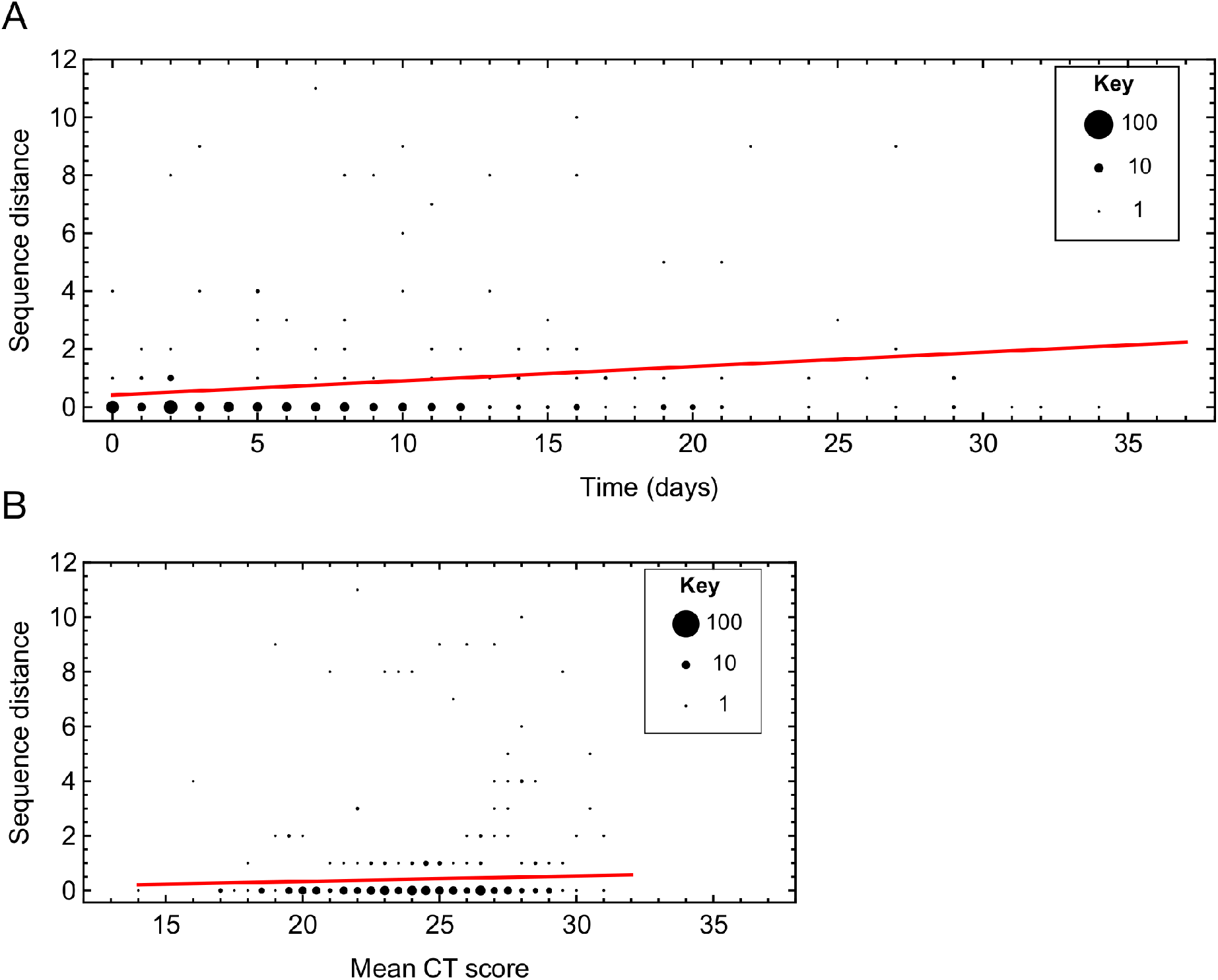
Analysis of Hamming distances between pairs of genome sequences collected from viral samples in the same host. Figures show projections through a multi-linear model fit to the data using a Poisson likelihood. **A.** Relationship between the Hamming distance and time between samples. The line shows the fit to the data at the mean CT score. The size of a dot is proportional to the number of pairs with given parameters. **B.** Relationship between the Hamming distance and mean CT score of the two samples. The line shows the fit to the data calculated at zero time between samples.

**S2 Figure:**
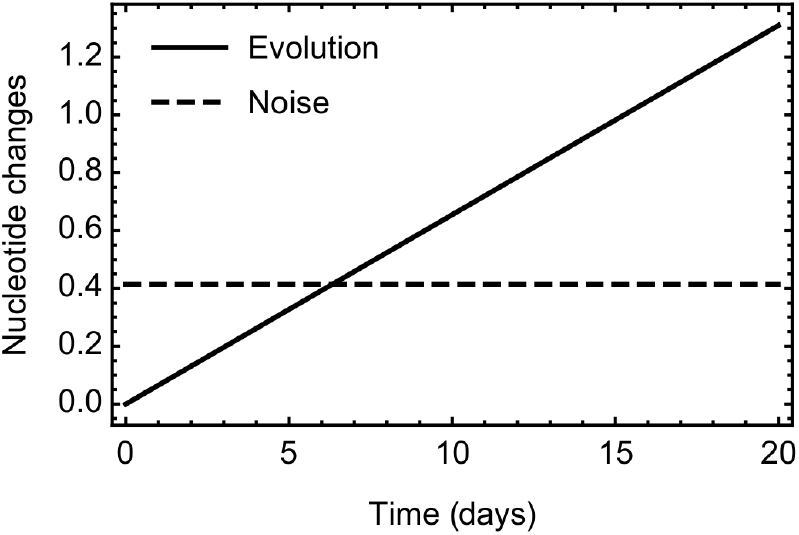
Comparison between the expected rate of SARS-CoV-2 evolution within our model, and the expected difference between two sequences caused by noise. The expected time between symptoms being reported from individuals in a transmission pair is 5.7 days, in which time the expected number of substitutions arising via evolution is 0.373. The expected number of differences between two genome sequences resulting from error was estimated as 0.414.

**S3 Figure:**
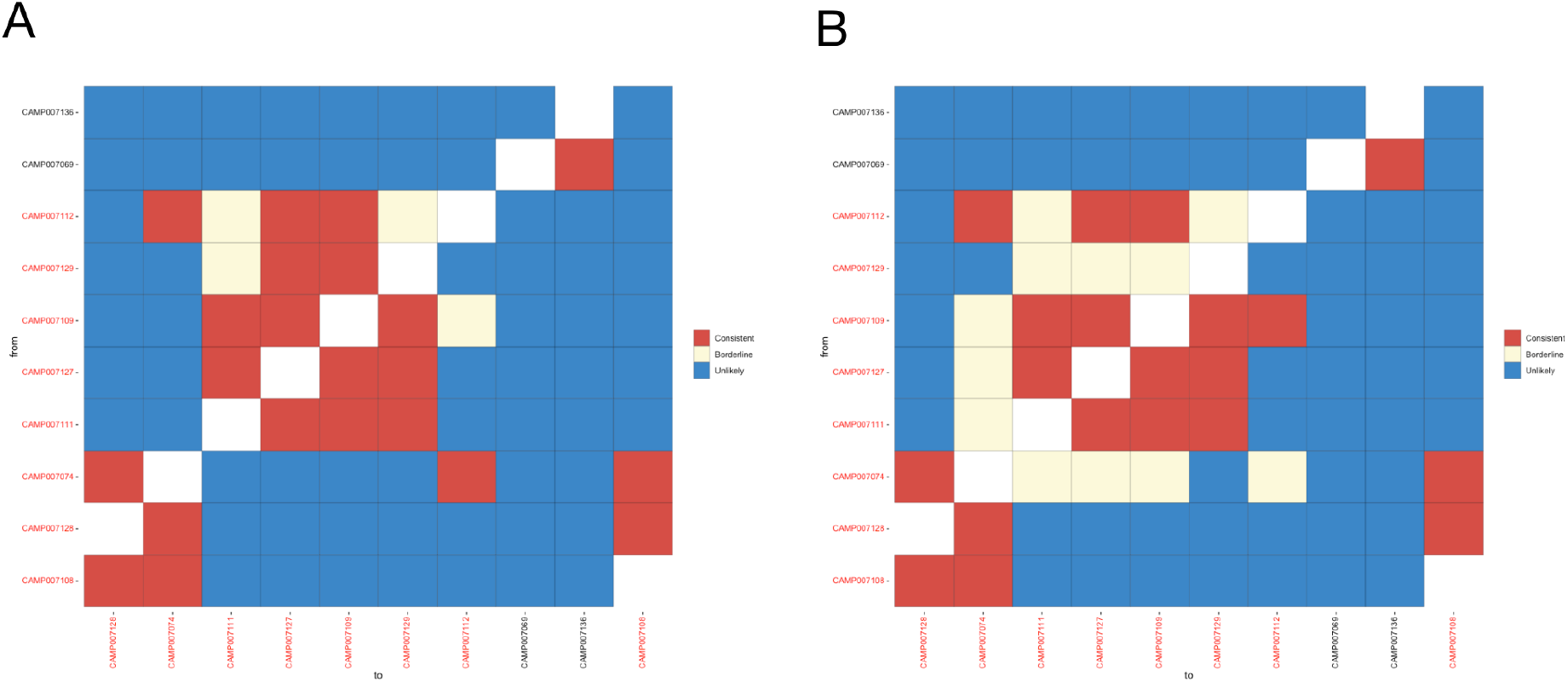
Sensitivity of our results to changes in the noise parameter. A. Inferences of potential transmission events for ward X given a noise parameter of zero. B. Inferences of potential transmission events for ward X given a noise parameter double that inferred from our data.

**S4 Figure:**
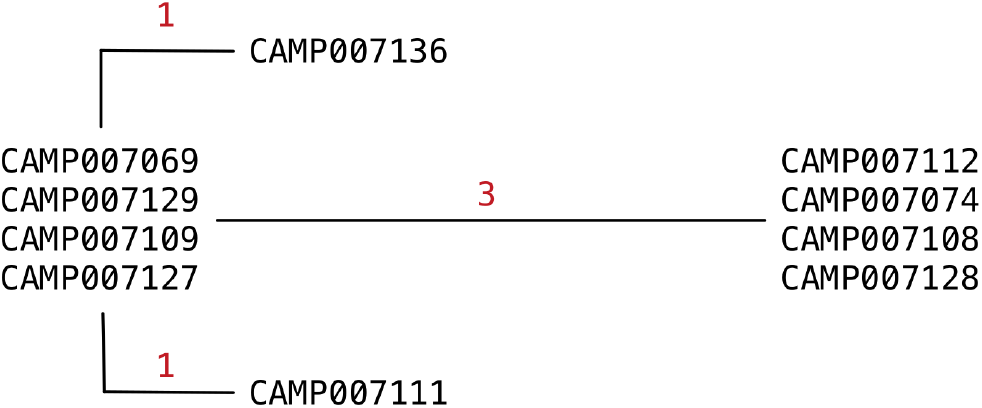
Phylogenetic relationship between sequences collected from individuals on ward X. The tree was constructed manually using a maximum parsimony method. Red digits indicate the number of substitutions between sequences from each individual

**S5 Figure:**
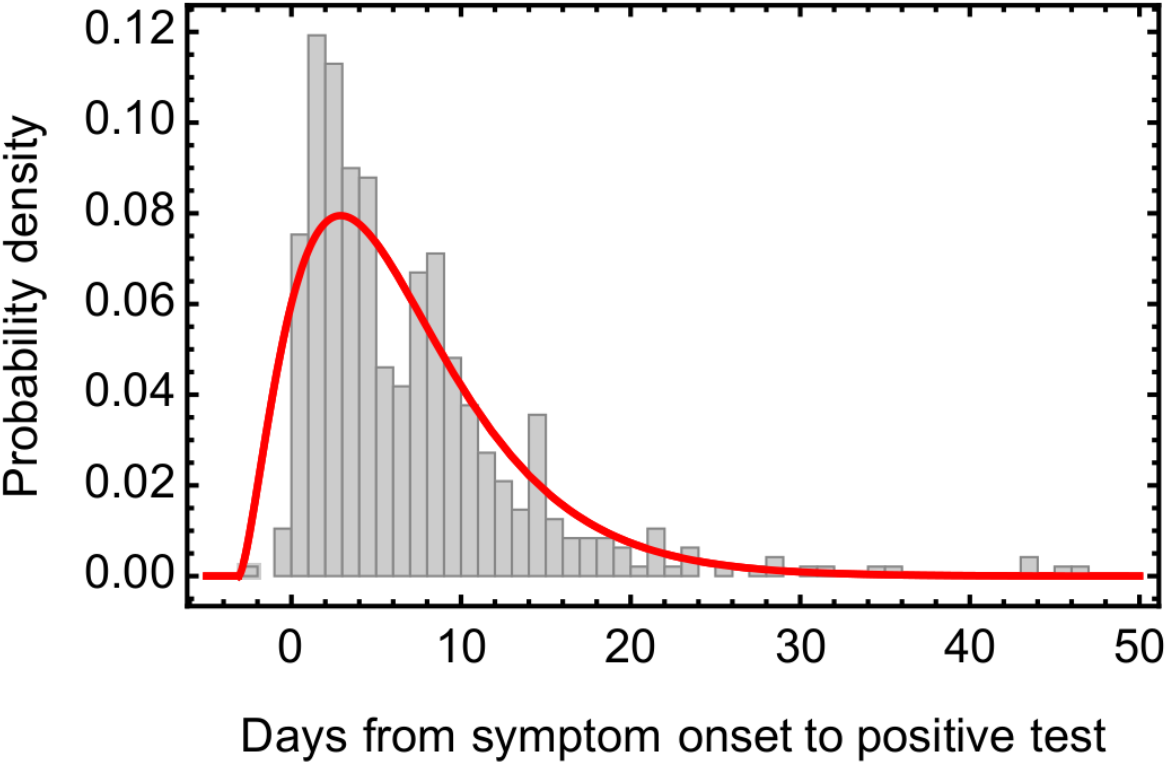
Raw data (bars) and inferred model (red line) describing the distribution of the time between the onset of symptoms and receiving a positive test. This model was used to impute? equivalent symptom onset dates for individuals who were asymptomatic or for whom no data on symptom onset date were available.

**S1 Table:**
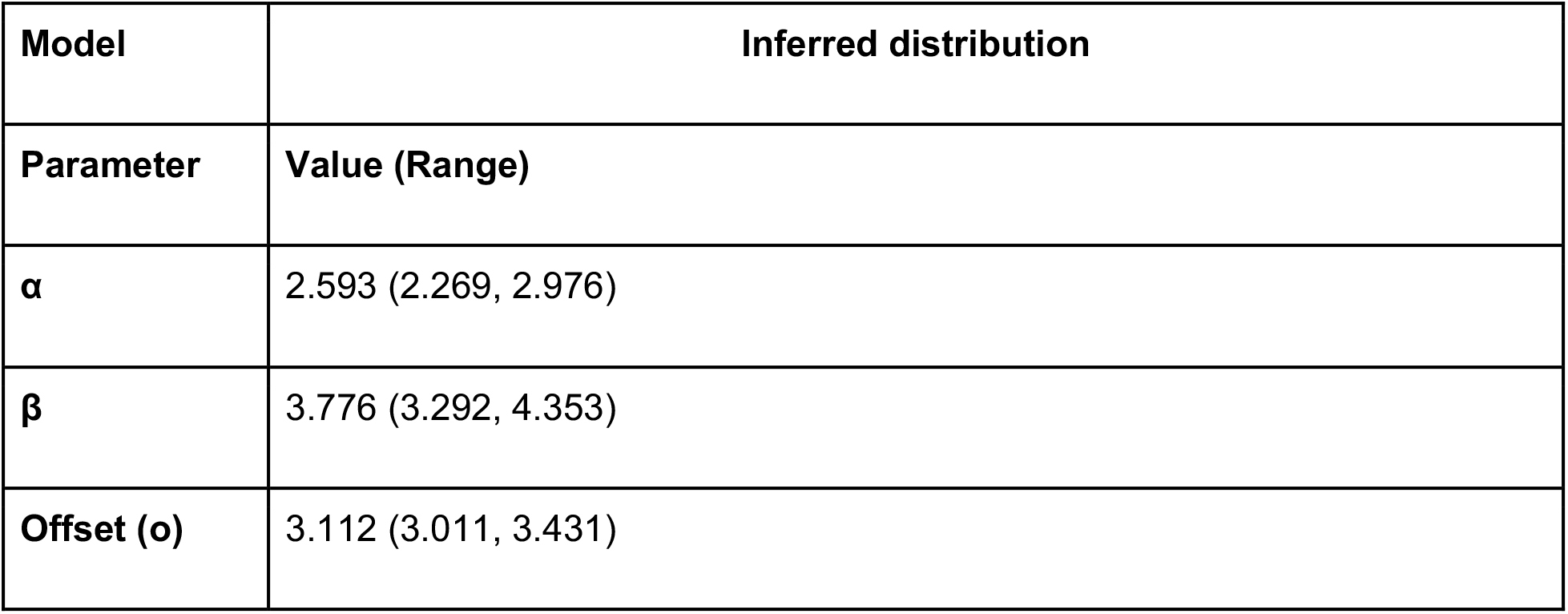
Parameters for the offset gamma distribution fitted to data describing intervals between times of reporting symptoms and positive test results. Inferred values were generated using maximum likelihood; the range describes a window of size two likelihood units from the maximum.

**S2 Table:** Error models fitted to Hamming distance data. A model incorporating dependence upon the time between samples and upon viral load gave the best fit to the data. The error parameter used was calculated at zero time between samples and at the mean viral load.

| Model | Parameters |  |  | BIC |
| --- | --- | --- | --- | --- |
| | Constant ($\lambda$) | Time dependence ($\gamma$) | CT dependence ($\alpha$) | |
| Constant error | 0.842 |  |  | 887.3 |
| Time-dependent | 0.385 | 0.0525 |  | 858.5 |
| Time and CT<br>dependent | -0.0692 | 0.0492 | 0.0200 | 857.8 |

### S1 Text: Further methodological details

In the main text we stated that:

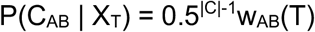

To derive this result we note that, if it is observed that C_AB_(T)=0, transmission cannot occur at time T, so that P(C_AB_|X_T_)=0. If it is observed that C_AB_(T)=1, we next consider the element C_AB_(t) of C_AB_ for a time t≠T. If C_AB_(t) is observed, we apply our approach to contact patterns, assuming that P(C_AB_(t)=1|X_T_) = P(C_AB_(t)=0|X_T_) = 0.5, such that the probability of this observation is 0.5. If C_AB_(t) is not observed, its probability is obtained by integration. We have that P(C_AB_(t) | X_T_) = w_AB_(t)* 0.5 + (1 - w_AB_(t)) * 0.5 = 0.5. Hence if C_AB_(T)=1, the probability P(C_AB_ | X_T_) of the whole contact vector is equal to 0.5^|C|-1^. Finally, we consider the case in which C_AB_(T) is missing data. Integrating over the missing value, we have that

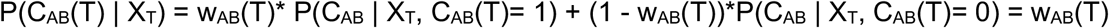

Applying again the reasoning above, this gives us the result P(C_AB_ | X_T_) = 0.5^|C|-1^w_AB_(T). As we defined w_AB_(T) = C_AB_(T) when C_AB_(T) was observed, we thus have that P(C_AB_ | X_T_) = 0.5^|C|-1^w_AB_(T) in every case.

### S2 Text: GISAID identifiers for sequences used in this study

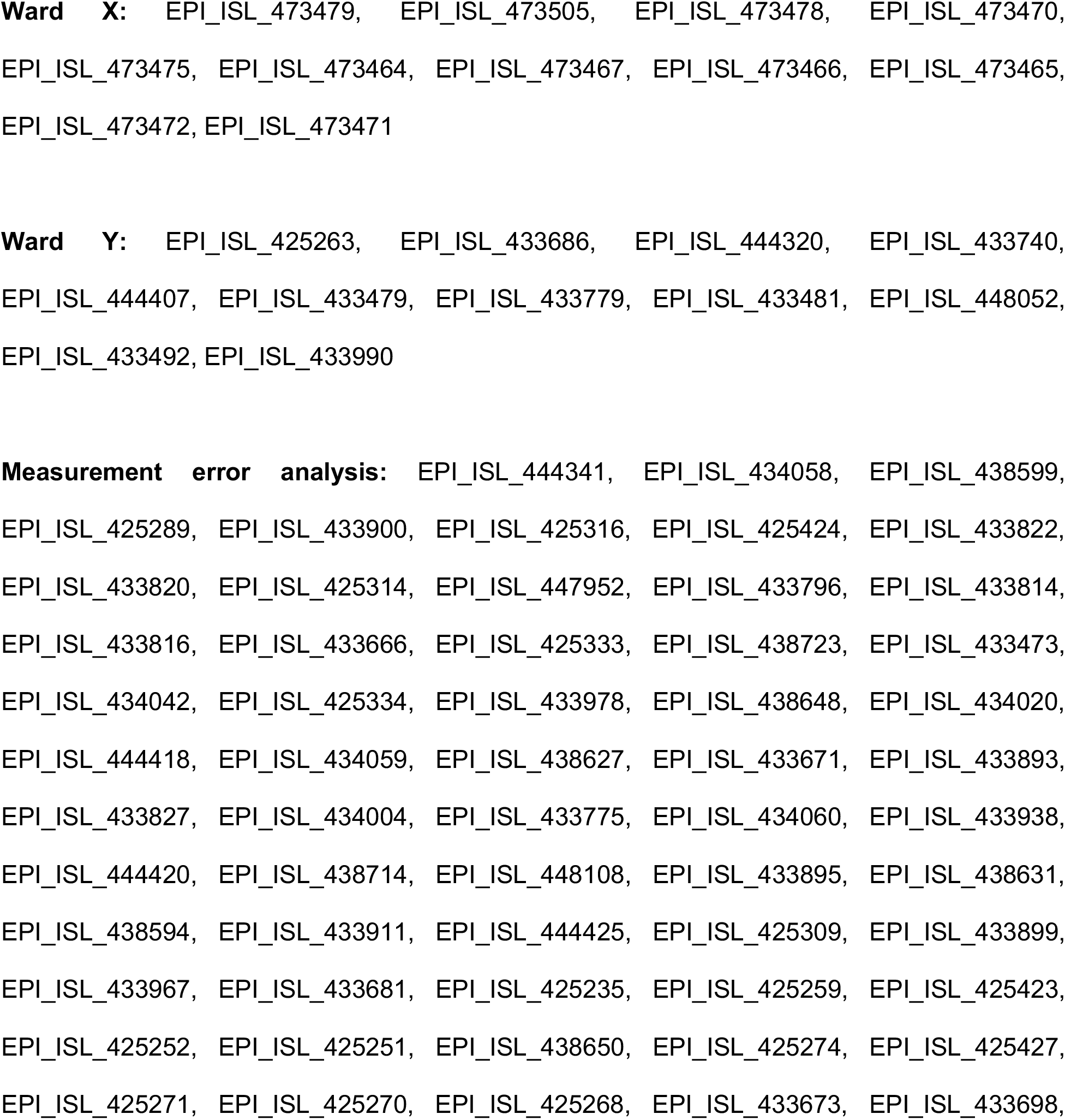
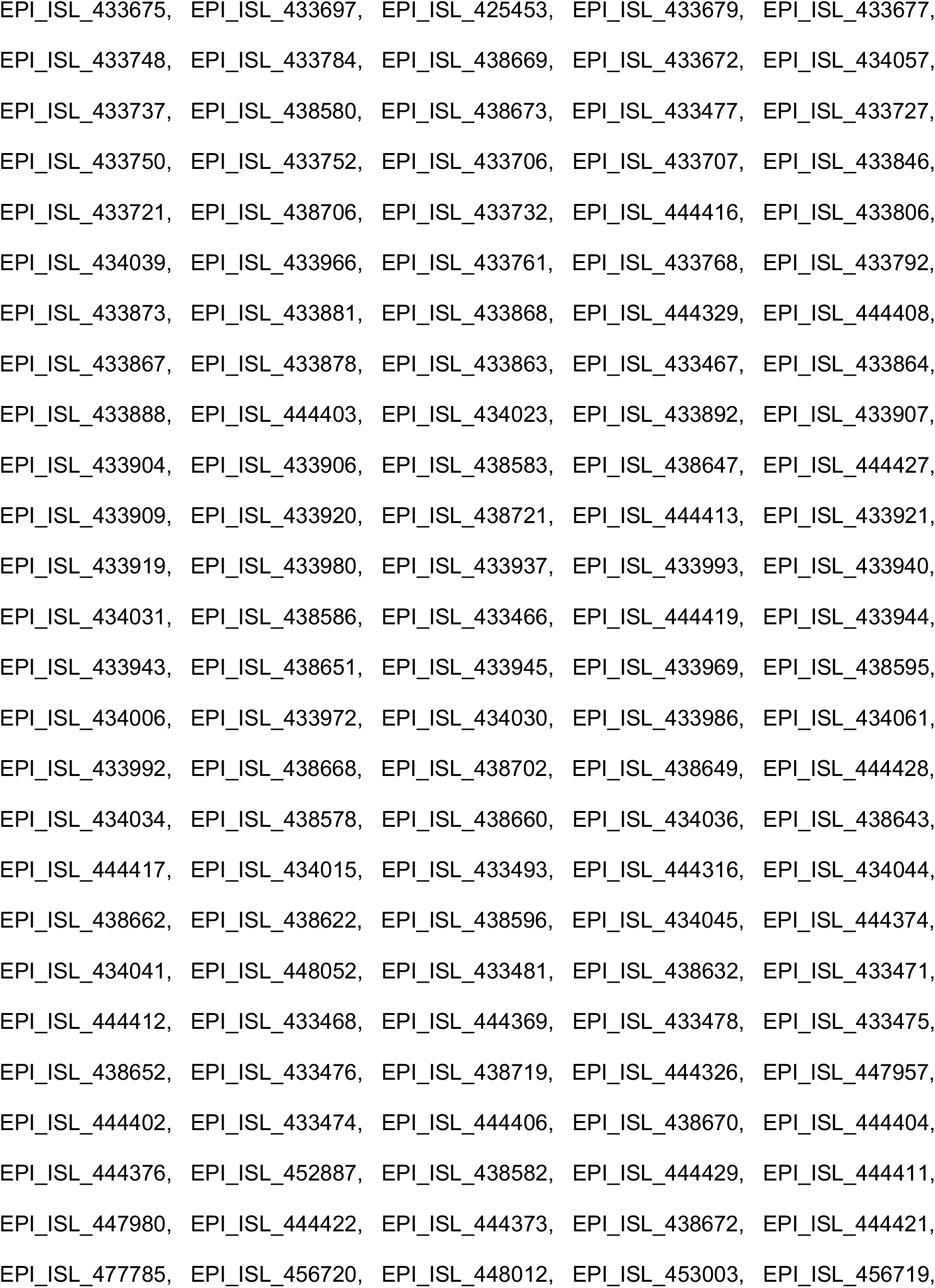
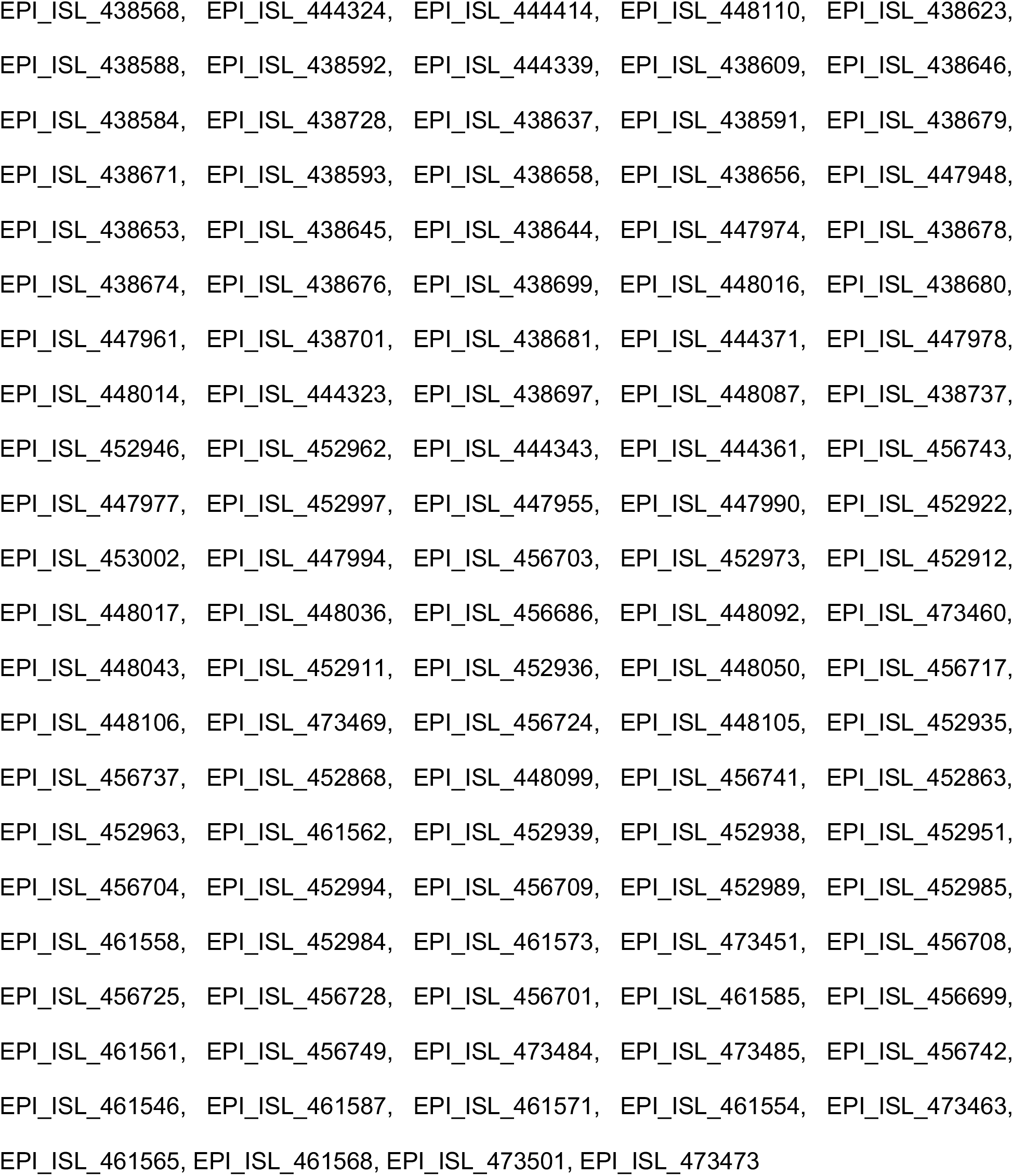

## Author contributions

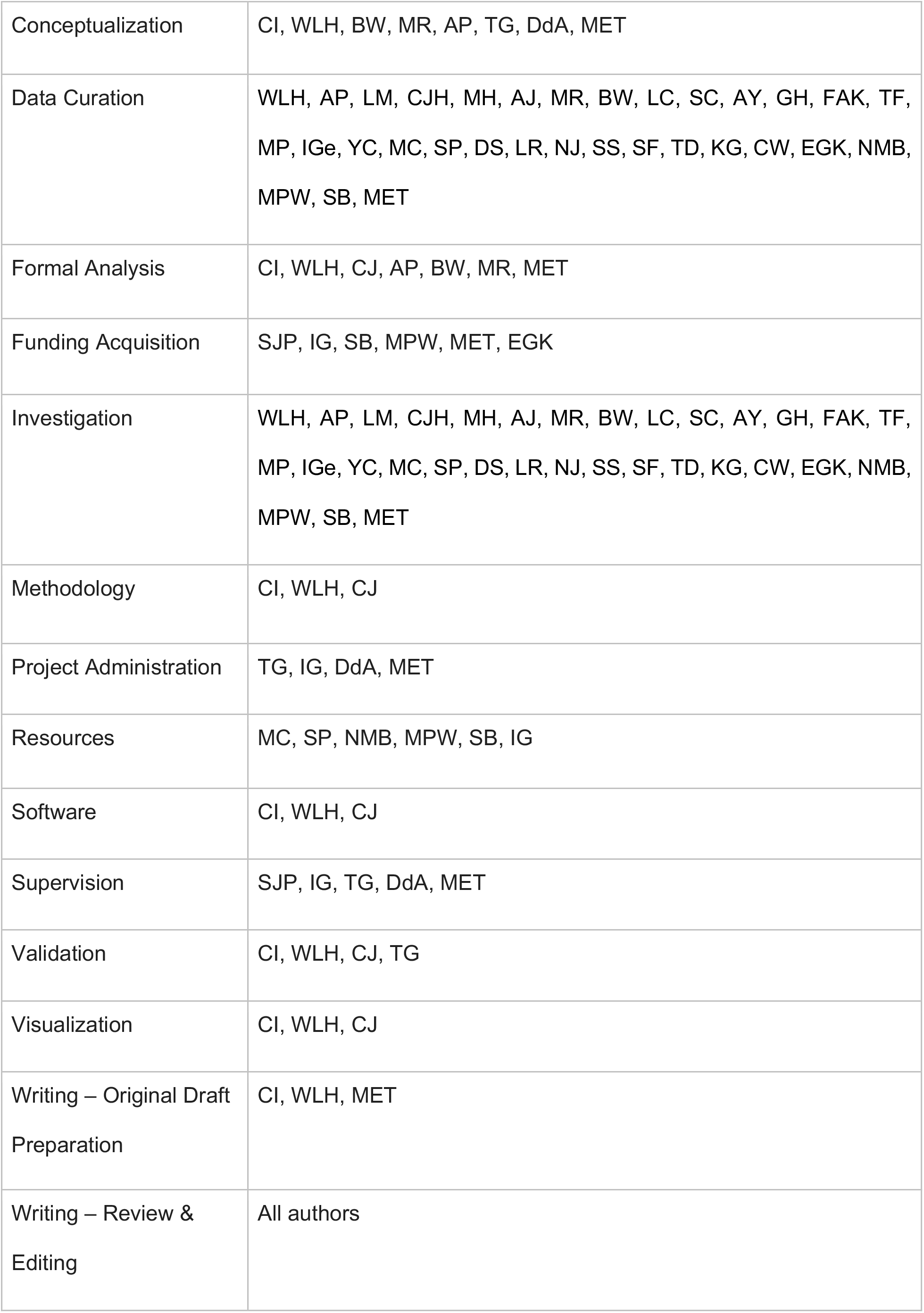

## Acknowledgements

This work was funded by COG-UK, which is supported by funding from the Medical Research Council (MRC) part of UK Research & Innovation (UKRI), the National Institute of Health Research (NIHR) and Genome Research Limited, operating as the Wellcome Sanger Institute; We also acknowledge the support from the Wellcome (Senior Clinical Fellowship to MPW (ref: 108070/Z/15/Z), Senior Research Fellowship to SB (ref: 215515/Z/19/Z), Senior Fellowship to IG (ref: 097997/Z/11/Z); Collaborative Grant to CJH (ref: 204870/Z/16/Z); the Academy of Medical Sciences & the Health Foundation (Clinician Scientist Fellowship to MET), the NIHR Cambridge Biomedical Research Centre (to BW, MET) and the NIHR Clinical Research Network Greenshoots award (to EGK). CJRI was supported by Deutsche Forschungsgemeinschaft (DFG) Grant SFB 1310. We acknowledge MRC funding (ref: MC_UU_00002/11).

## Availability of code

Our app is suitable for use with the R package and can be downloaded from http://github.com/chjackson/a2bcovid.

## Notes

### Competing Interest Statement

The authors have declared no competing interest.

### Summary of Updates

Methods section revised and clarified. Alteration to calling thresholds.

